# Genetic and morphological estimates of androgen exposure predict social deficits in multiple neurodevelopmental disorder cohorts

**DOI:** 10.1101/2020.08.03.20155671

**Authors:** Brooke G. McKenna, Yongchao Huang, Kévin Vervier, Dabney Hofamman, Mary Cafferata, Seima Al-Momani, Florencia Lowenthal, Angela Zhang, Jin-Young Koh, Savantha Thenuwara, Leo Brueggeman, Ethan Bahl, Tanner Koomar, Natalie Pottschmidt, Taylor Kalmus, Lucas Casten, Taylor R. Thomas, Jacob J. Michaelson

**Affiliations:** Department of Psychology, Emory University, Atlanta, USA; Department of Psychiatry, University of Iowa, Iowa City, USA; Host-Microbiota Interactions Laboratory, Wellcome Sanger Institute, Hinxton, UK; Department of Psychology, University of Nebraska, Omaha, USA; University of Washington School of Public Health, Seattle, USA; Molecular Otolaryngology and Renal Research Laboratories, University of Iowa, Iowa City, USA; Biomedical Science Department, Iowa State University, Ames, USA; Department of Psychology, Pennsylvania State University, State College, USA; Department of Psychology, University of Washington, Seattle, USA

**Author notes:** equal contribution (Brooke G. McKenna, Yongchao Huang).

## Abstract

**Background:** Neurodevelopmental disorders (NDDs) such as autism spectrum disorder (ASD) display a strong male bias. Androgen exposure is profoundly increased in typical male development, but it also varies within the sexes, and previous work has sought to connect morphological proxies of androgen exposure, including digit ratio and facial morphology, to neurodevelopmental outcomes. The results of these studies have been mixed and the relationships between androgen exposure and behavior remain unclear.

**Methods:** Here, we measured both digit ratio masculinity (DRM) and facial landmark masculinity (FLM) in the same neurodevelopmental cohort (N=763) and compared these proxies of androgen exposure to clinical and parent-reported features as well as polygenic risk scores.

**Results:** We found that FLM was significantly associated with NDD diagnosis (ASD, ADHD, ID; all *p* < 0.05), while DRM was not. When testing for association with parent-reported problems, we found that both FLM and DRM were positively associated with concerns about social behavior (*ρ* = 0.19, *p* = 0.004; *ρ* = 0.2, *p* = 0.004, respectively). Furthermore, we found evidence via polygenic risk scores (PRS) that DRM indexes masculinity via testosterone levels (*t* = 4.0, *p* = 8.8 × 10^−5^), while FLM indexes masculinity through a negative relationship with sex hormone binding globulin (SHBG) levels (*t* = −3.3, *p* = 0.001). Finally, using the SPARK cohort (N=9,419) we replicated the observed relationship between polygenic estimates of testosterone, SHBG, and social functioning (*t* = −2.3, *p* = 0.02, and *t* = 4.2, *p* = 3.2 × 10^−5^ for testosterone and SHBG, respectively). Remarkably, when considered over the extremes of each variable, these quantitative sex effects on social functioning were comparable to the effect of binary sex itself (binary male: −0.22 ± 0.05; testosterone: −0.35 ± 0.15 from 0.1%-ile to 99.9%-ile; SHBG: 0.64 ± 0.15 from 0.1%-ile to 99.9%-ile).

**Conclusions:** These findings and their replication in the large SPARK cohort lend support to the hypothesis that increasing net androgen exposure diminishes capacity for social functioning in both males and females.

## 1 Introduction

Most neurodevelopmental disorders (NDDs) present more often in males than females [1], and autism spectrum disorder (ASD) in particular shows a striking sex bias with a 4:1 male-to-female ratio. However, despite the evidence that male sex – and masculinity more generally, as posited by the ‘extreme male brain’ theory [2] – is a strong risk factor for ASD, the cause of this sex bias is not yet well understood. The ‘extreme male brain’ theory suggests that the male sex bias may reflect the role of male sex hormones (i.e., androgens) in the etiology of ASD; specifically, risk for ASD may stem from increased androgen (e.g., testosterone) exposure during critical periods of fetal development, as the brain undergoes substantial organization and sexual differentiation [3] [4]. Indeed, numerous studies have found that greater prenatal androgen exposure is associated with ASD and ASD-related traits [5] [6] [7] [8], although other studies have observed no relationship [9] [10]. Postnatal studies utilizing clinical and community samples of adults have also identified links between testosterone levels and ASD-related traits [11] [12], although postnatal studies of children show mixed results [13] [14]. Interestingly, despite evidence from many of these studies that greater testosterone is associated with phenotypes that characterize neurodevelopmental disorders more broadly – for instance, poor language abilities [15] [16] – the majority of studies that examine androgen exposure focus on ASD. Given the mixed evidence and dearth of studies examining NDDs outside of ASD, a better understanding of whether and how testosterone exposure may contribute to the male bias observed across all NDDs is warranted [8].

The links between androgen exposure and neurodevelopment have primarily been established via cross-sectional studies that measure testosterone levels at a single time point during perinatal (e.g., amniotic fluid, umbilical cord blood) or postnatal (e.g., blood serum, saliva) development. Beyond the practical challenges of measuring testosterone via biological samples, particularly during the perinatal period, these measures provide only a snapshot of androgen exposure at the time of collection, rather than capturing an individual’s cumulative exposure across development. To overcome the limitations of measuring testosterone levels directly, morphological and genetic measures are increasingly being leveraged to indirectly assess androgen-related risk. Digit ratio, or the ratio in length of the second and fourth digit (i.e, 2D:4D ratio), is a sexually dimorphic feature that is lower in males than in females [17] and has been linked to fetal testosterone exposure [18]. Numerous studies indicate that individuals with ASD exhibit lower (i.e., more masculine) digit ratios than their typically-developing counterparts [19] [20] [21] [22] [23], and evidence suggests this may be true for other NDDs as well [24] [25] [26]. However, a number of recent studies have failed to replicate the association between fetal testosterone exposure and digit ratio [27] [28], indicating that digit ratio may be a useful biomarker of NDDs but its biological underpinnings require further clarification.

Facial masculinity has also been proposed as a potential indicator of fetal testosterone and NDD-related risk. A twin study found that females with a male twin, who were presumably exposed to greater fetal testosterone levels, exhibited greater facial masculinity than those with a female twin [29] and another study showed that greater umbilical cord testosterone levels were associated with greater adult facial masculinity in both sexes [30]. Moreover, recent studies have found that facial masculinity predicts ASD traits in a variety of populations including children with ASD [31], siblings of individuals with ASD [32], as well as non-clinical samples [33] [34]. However, these studies provide inconsistent support for the ‘extreme male brain’ theory [35] [36]. While some studies found that greater masculinity was associated with greater ASD traits in both sexes, others suggested that androgynous features may confer greater risk. For example, one study found that female subjects with ASD demonstrated more masculine faces than their typically-developing peers, but males with ASD exhibited *less* masculine faces than their peers [36]. Given that these early findings are mixed, and that a dearth of studies has examined facial masculinity in the context of other NDDs, it remains unclear whether facial masculinity can yield insight into NDDs and, if so, how it may reflect androgen-related risk.

Running parallel to morphological proxies of androgen exposure, genetic studies of NDDs can yield insight into the hypothesized role of androgens in the etiology of NDDs. While large-scale studies have so far not identified individual genomic loci that strongly implicate sex hormone pathways as a major risk factor, some smaller studies have yielded clues worthy of further investigation. For example, one study (*n* = 174) found that three genes (the estrogen receptor *ESR2* and two steroid enzymes, *CYP11B1* and *CYP17A1*) related to the synthesis, transport, or metabolism of sex hormones were associated with ASD-related traits in non-clinical samples [37]. Another study found that *CYP19A1*, a steroid enzyme that contributes to sexual differentiation of the brain, was associated with dyslexia, a form of language impairment [38]. Yet another (*n* = 1,171) identified three genes – that encoded an estrogen receptor (*ESR1*), steroid enzyme (*SRD5A2*), and sex-hormone binding globulin (*SHBG*) – as conferring risk for ASD-related traits in males [39], although replication in a larger sample (*n* = 10,654) found that only variation in *SHBG* emerged as a significant predictor [40]. Interestingly, levels of SHBG are inversely associated with levels of active (i.e., “free”) testosterone. Testosterone becomes inactive when it is bound to SHBG, and studies show that low levels of SHBG are found in individuals with excessive androgen activity [41]. These findings suggest that SHBG and other hormone-binding proteins may play an important role in the etiology of ASD through regulation of testosterone exposure. Moreover, given evidence that SHBG and free testosterone levels have been linked to attention-deficit/hyperactivity disorder (ADHD) [42] and that genome-wide studies have identified significant genetic overlap between ASD and other NDDs [43] [44], it is possible that variation in SHBG and other testosterone regulators may contribute to NDDs more broadly. Further investigation of these potential mechanisms is needed.

Together, these previous studies point to several important yet unanswered questions. First, which morphological proxies of androgen exposure are most predictive of NDD diagnosis? Second, what specific behavioral and cognitive phenotypes are most related to these proxies of androgen exposure? Third, do morphological proxies such as digit ratio and facial morphology capture the same or different molecular aspects of masculinization, and how do these mechanisms relate to neurodevelopmental risk? Finally, how do quantitative effects related to androgen exposure in both sexes compare to the binary sex effect of being a Y-chromosomal male? The answers to these questions will deliver vital insight into the nature of male bias in NDDs. Toward this end, we carried out an investigation in two steps (Figure 1). First, we assembled a neurodevelopmental cohort (N=763) that includes 1) genome-wide genotypes, 2) measures of digit ratio masculinity (DRM), 3) measures of facial landmark masculinity (FLM) derived by machine learning, and 4) extensive clinical and parent-reported phenotypic information. This is the first study to investigate genetic factors associated with both digital and facial masculinity and their connection to neurodevelopmental outcomes. Second, to ensure the robustness of findings generated by investigation of our discovery cohort, we employed SPARK, a large genetic study of autism (N=9,419 in this analysis), as a means for testing the generalization of our observations.

**Figure 1.**
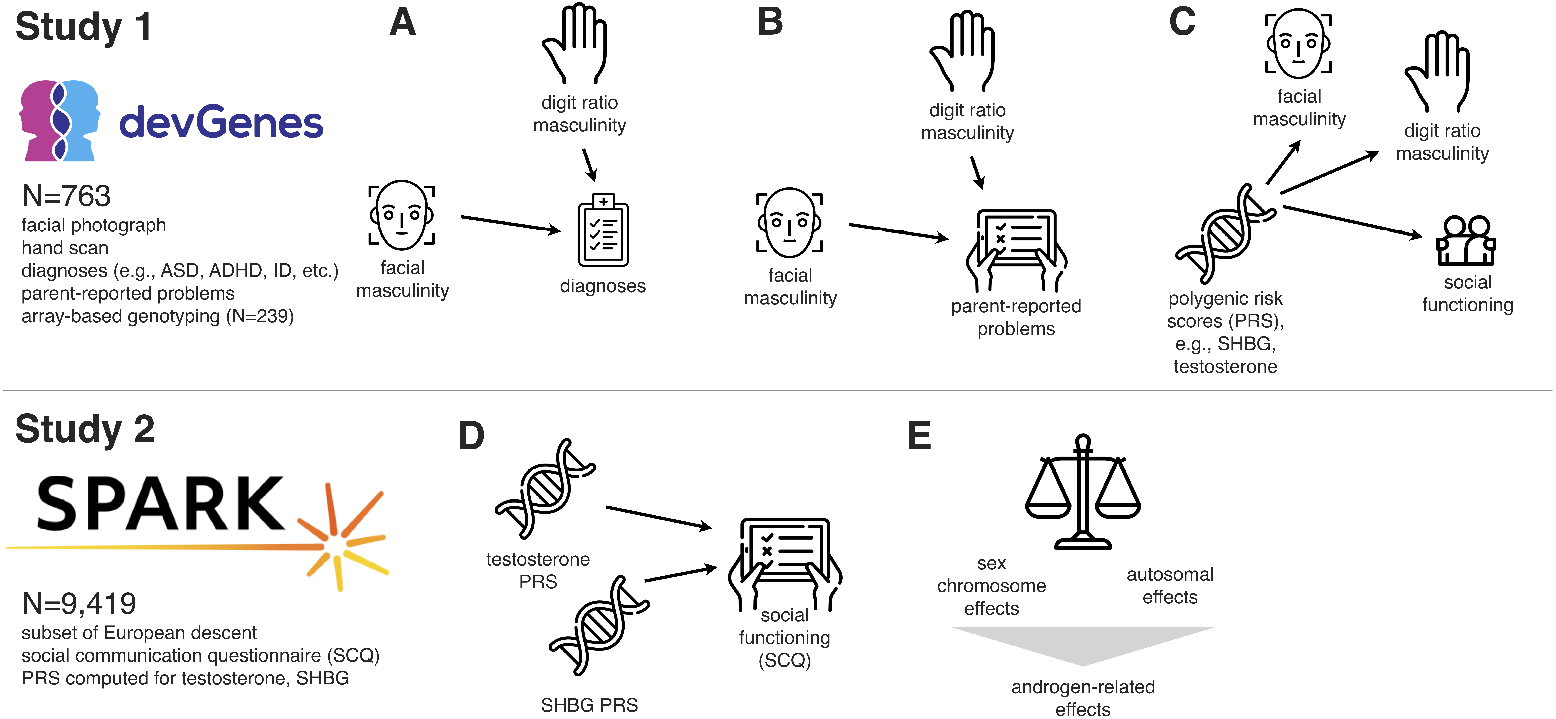
Study overview. Study 1 (devGenes, N=763) investigated computer-generated proxies for masculinization that are evident in facial photographs and the 2D:4D ratio (hand scans), and the relationship of these masculinity measures to diagnoses (A), parent-reported problems (B), and polygenic risk scores (C). These investigations suggested involvement of androgen-related mechanisms in social functioning, a hypothesis we investigated further in study 2 (SPARK, N=9,419). In SPARK, we computed polygenic risk scores for testosterone levels and sex hormone binding globulin (SHBG) levels, and tested those for association with latent factors in the social communication questionnaire (SCQ) (D). This analysis revealed that binary sex effects (male or female) on these factor scores and items were comparable in magnitude to the effect of the polygenic risk scores, which are derived entirely from autosomal genotypes (E).

## 2 Methods

### 2.1 Sample: devGenes

devGenes is a neurodevelopmental registry drawing participants largely from the midwestern United States and Iowa in particular. From 2015 to 2020, individuals of any age with a diagnosis of attention-deficit/hyperactivity disorder (ADHD), autism spectrum disorder (ASD), intellectual disability (ID), language impairment, and/or pediatric epilepsy (including individuals with a syndromic form of NDD [n=15; 2.0% of total sample, see Supplementary Table 1]) were recruited to participate in a genetic study of neurodevelopmental disorders. Biological relatives, regardless of age or affected status, were invited to participate as well. As such, the devGenes sample is enriched for individuals with NDDs but includes participants across a broad range of diagnoses and ages (i.e., 2 to 80 years; Table 1).

**Table 1.** Demographics (devGenes)

| Name | Male (N=381) |  | Female (N=382) |  |
| --- | --- | --- | --- | --- |
| | N / Mean | Percent / ± SD | N / Mean | Percent / ± SD |
| age | 21.31 | 17.45 | 30 | 18.15 |
| Caucasian | 264 | 83.3% | 273 | 85.6% |
| ASD | 135 | 35.40% | 46 | 12.00% |
| ADHD | 96 | 25.20% | 52 | 13.60% |
| ID | 97 | 25.50% | 36 | 9.40% |
| epilepsy | 12 | 3.10% | 8 | 2.10% |
| language impairment | 112 | 29.40% | 44 | 11.50% |
| depression | 19 | 5.00% | 36 | 9.40% |
| bipolar disorder | 4 | 1.00% | 9 | 2.40% |
| anxiety | 28 | 7.30% | 34 | 8.90% |
| unaffected | 165 | 43.30% | 253 | 66.20% |
| genetic syndromes | 10 | 66.67% | 5 | 33.33% |

Participants were informed of the study through a combination of public flyers, personal letters, follow-up phone calls, and local clinicians or community leaders. Participants met with a trained member of the research team for a single 60-minute visit in individuals’ homes, at independent and state-funded clinics, and in research laboratories at the University of Iowa. Study procedures were approved by the University of Iowa’s Institutional Review Board, and informed consent was obtained for each participant (IRB 201505743).

### 2.2 Sample: SPARK

SPARK is a nationwide (U.S.) genetic study of autism funded by the Simons Foundation [45]. Genetic and self- and parent-reported demographic and phenotypic data are made available to researchers through https://base.sfari.org. We used the available genome-wide common variant genotyping data (described below) as well as results from the Social Communication Questionnaire (SCQ) [46], SPARK version 4 phenotype release.

### 2.3 Diagnostic and parent-report data

Participants in devGenes or their legal guardians were asked to complete a detailed survey regarding the participant’s demographics, developmental history, neurodevelopmental diagnoses (i.e., ADHD, ASD, language impairment (including dyslexia), and pediatric epilepsy), and lifetime mood/anxiety diagnoses (i.e., major depressive disorder, bipolar disorder, and generalized anxiety). A total of *N* = 239 participants had parent-report data from 38 items in total (mean item missingness: 1.5%). Parent-report survey items were arranged by thematic group and their reliability assessed by Cronbach’s alpha (Supplementary Table 2). After survey completion, reported medical history and diagnoses were confirmed where possible by a member of the research team through examination of participants’ University of Iowa Hospital and Clinics medical records. All devGenes data were stored in a REDCap database [47] [48]. devGenes participants were classified as “affected” for analysis purposes if they reported any of the following diagnoses: ADHD, autism, intellectual disability, language impairment, or pediatric epilepsy. Participants were classified as “unaffected” if they did not endorse any of the five NDD diagnoses, even they reported a mood/anxiety diagnosis. In order to capture overall diagnostic burden, the total number of reported diagnoses (including both NDDs and mood/anxiety disorders) were summed for each participant, with levels of 0, 1, 2, and 3 or more. Basic individual-level data for devGenes participants is included in Supplementary Table 3, and we have made code available at https://research-git.uiowa.edu/michaelson-lab-public/facial-masculinity-2021 to reproduce the analyses presented in figures 2-4.

**Table 2.**
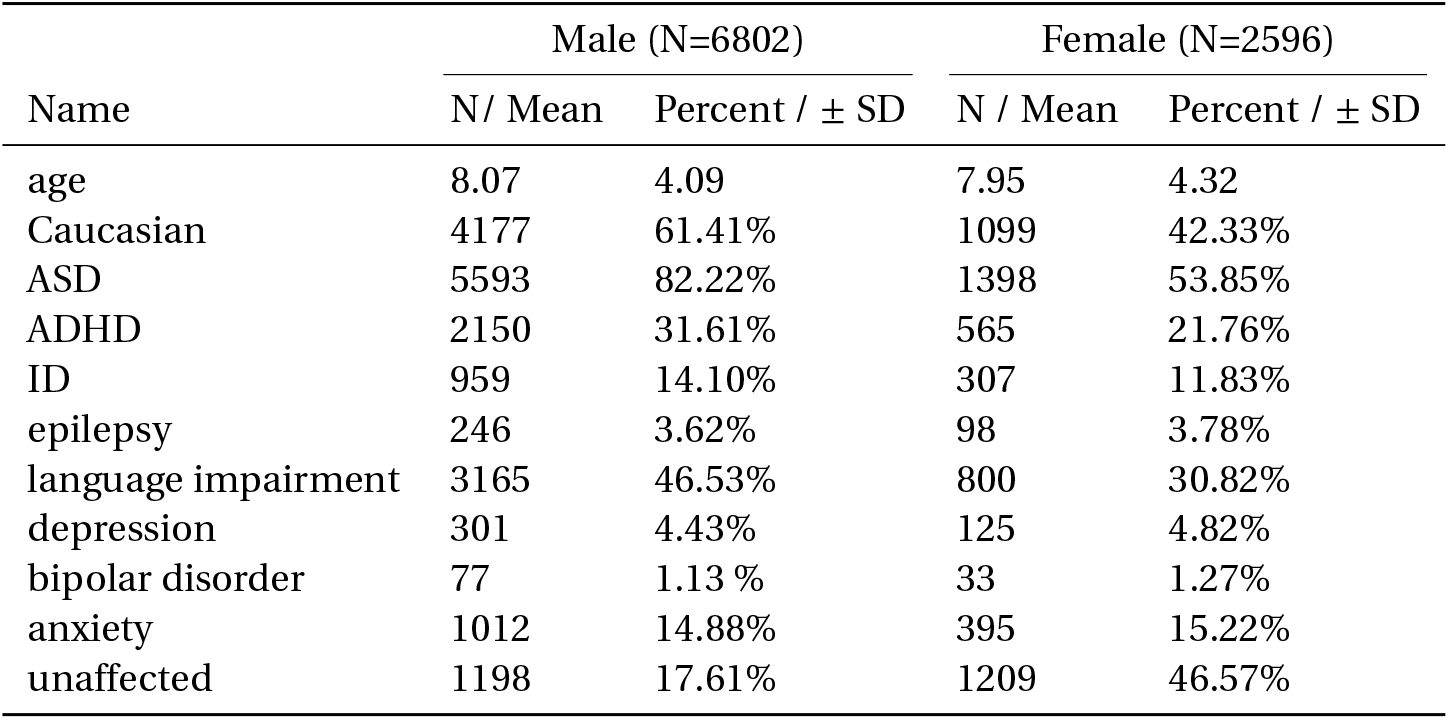
Demographics (SPARK)

**Figure 2.**
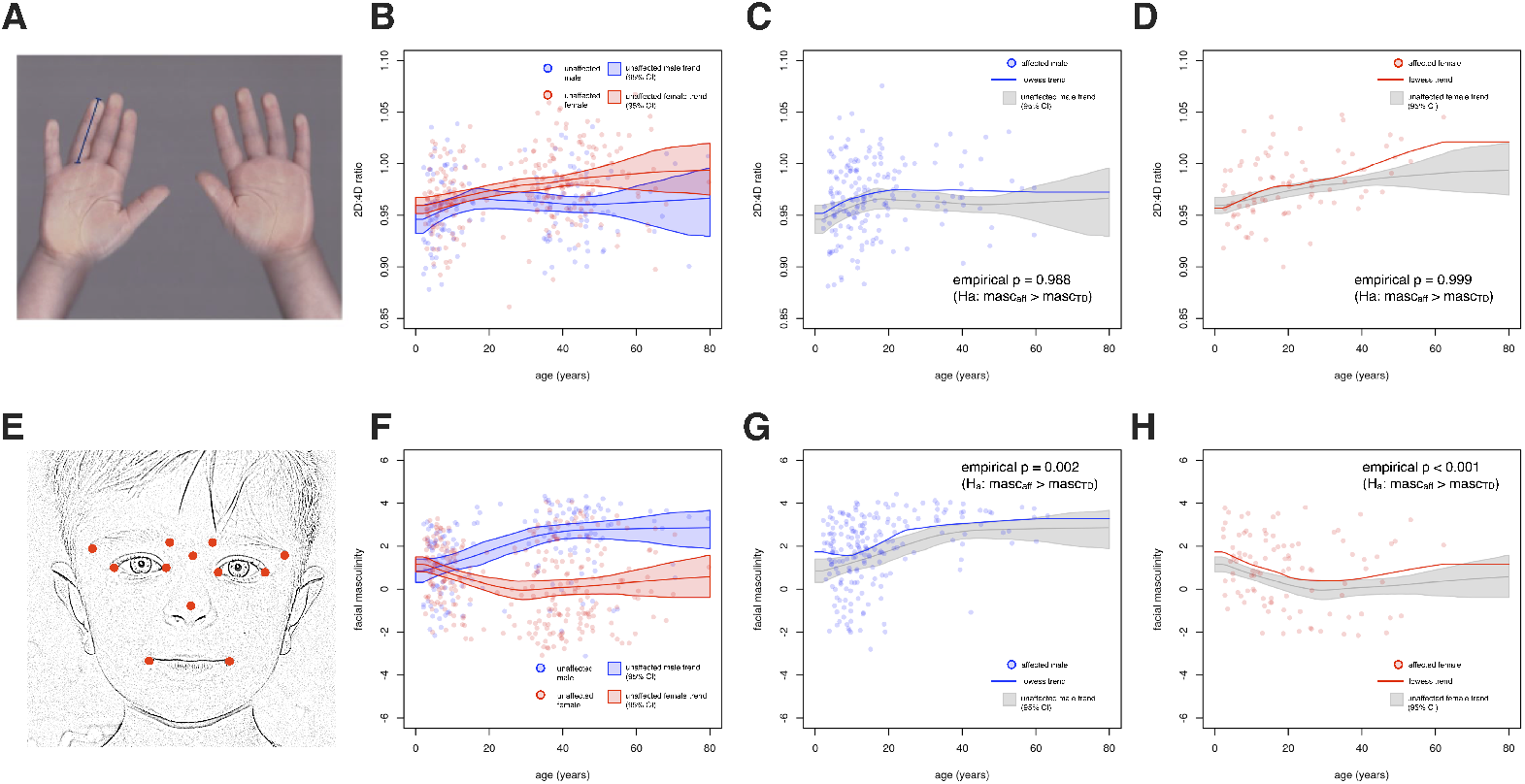
Masculinity as measured by digit ratio and facial landmarks. (A) Hand scans from devGenes participants were used to measure digit lengths and subsequently the ratio between the index and ring fingers (2D:4D ratio). (B) Cross-sectional data were collected from individuals spanning from age 2 to age 80. The 95% interval of the lowess trend of 2D:4D ratio vs. age for typically developing males and females is shown.(C, D) The 2D:4D ratio of affected males and affected females is shown with respect to the 95% confidence interval for their typically developing counterparts, with accompanying empirical p values. (E) Facial photographs from devGenes participants were used to calculate facial masculinity using a Random Forest classifier. (F) The lowess trends and 95% confidence interval for typically developing males and females is shown. (G, H) The facial masculinity of affected males and affected females is shown with respect to the 95% confidence interval for their typically developing counterparts, with empirical p values.

### 2.4 Masculinity measures

#### Hand Scans / Digit Ratio Masculinity (DRM)

Hand scans from devGenes participants were acquired using a flatbed image scanner operated by a trained research assistant. Participants were instructed to place both hands on the scanner, fingers spread and fully extended, to obtain a single scan of all ten fingers. The lengths of index fingers (i.e., second digits [2D]) and ring fingers (i.e., fourth digits [4D]) on both hands were measured manually using Image-J [49] (Fig. 2A).

Digit lengths were then corrected for rater and scanner effects with a linear model before being used to calculate left and right 2D:4D digit ratio. Using unaffected males and females, a lowess curve was fit that defined the trend, for each sex separately, of 2D:4D ratio with respect to age. To determine the 95% confidence interval of this trend, we performed bootstrap resampling 1,000 times, each time recording the lowess curve (Fig. 2B). The mean curve of these bootstrap samples was then used as the point of reference, and all individual data points for either males or females were transformed to Z-statistics using the mean curve for that sex, and point estimates of the standard deviation. Finally, we multiplied these Z-statistics by −1 so that increasingly positive values corresponded to increasing digit ratio masculinity. These Z-statistics (N=664) were used in all subsequent analyses as “digit ratio masculinity” (DRM).

#### Two-Dimensional Facial Photography / Facial Landmark Masculinity (FLM)

Facial images from devGenes participants were acquired using a Nikon D3000 camera operated by a trained research assistant. Participants were instructed to look directly at the camera and maintain a neutral facial expression with closed lips (Fig. 2E).

Facial masculinity was assessed through computational analysis of the two-dimensional facial photographs. Coordinates (x, y) of 68 points(see Supplementary Fig.1A) were extracted using dlib [50], a computer vision library that contains algorithms for machine learning, computer vision, and image processing, including facial landmark detection. We validated this approach by comparing it with manual annotation of a provisional set of 17 facial landmarks on a subset of 99 pictures, in which the landmarks were manually selected to represent the overall facial structure with less redundancy. Results from this analysis indicated significant concordance overlap of coordinates between manual annotations and `dlib` (average Spearman correlation: 0.962; 95%CI [0.959,0.966]). For the present study, we used a subset of 12 coordinates (Fig. 2A), given the redundancy of landmarks within very close proximity and large variation of points around the jaw and chin region (Spearman correlation < 0.9). Landmark coordinates were normalized to a [0,1] interval in both x and y directions, as facial photographs tend to have different dimensions. The Euclidean distance between landmark pairs coordinates was then computed and used to provide 66 distance features [51].

A Random Forest classifier [52] [53] was then trained to discriminate males from females using these 66 features, as well as age in years (67 features total). Stratified sampling (via the `strata` and `sampsize` arguments to the `randomForest` function [54]) was performed so that all eight combinations of sex, age (dichotomized at 18 years), and affected status were approximately balanced at N=75 for each of the eight groups, for each tree of the Random Forest. The `ntree` parameter was set to 5,000 and `mtry` was set to 2 to reduce the greediness of the learning and increase diversity among the trees. Out-of bag estimates of misclassification showed a 38 % misclassification rate for females and a 33 % misclassification rate for males. The proportion of votes for class: male was used as the measure of masculinity. This vote proportion was transformed with a logistic function that put the midpoint of the curve at 0.4, which represents the vote proportion threshold that maximized the out-of-bag sensitivity and specificity, as determined by the F measure. These recalibrated class probabilities were then mapped onto a standard normal distribution. Next, to both observe and correct for the effects of age and sex on this masculinity estimate, we used unaffected males and females to fit a lowess curve that defined the trend, for each sex separately, of facial masculinity with respect to age. To determine the 95% confidence interval of this trend, we performed bootstrap resampling 1,000 times, each time recording the lowess curve (Fig. 2F). The mean curve of these bootstrap samples was then used as the point of reference, and all individual data points for either males or females were transformed to Z-statistics using the mean curve for that sex, and point estimates of the standard deviation. These Z-statistics (N=747) were used in all subsequent analyses as “facial landmark masculinity” (FLM). Individual-level FLM and DRM measures are included in Supplementary Table 3.

### 2.5 Genotyping and imputation (devGenes)

DNA was extracted from saliva using the Autogen QuickGene-610L kit (www.autogen.com, Catalog #FK-DBLC). The DNA quantity was assessed using the Qubit dsDNA High Sensitivity assay kit (www.thermofisher.com, Catalog #Q32854). The mean DNA concentration was 65.5 ng\µL. The DNA quality was assessed using 1% agarose gels. All samples were genotyped using Illumina Infinium PsychArray microarrays in two batches, the first using Illumina PsychArray v1.1, the second using PsychArray v1.3.

SNPs were mapped to the hg19 reference of the human genome. All Quality Control (QC) steps carried out with PLINK [55] and R [56], based on QC process described in [57]. R was used for calculating standard deviations from heterozygosity and population outliers. All other QC was carried out in PLINK. Samples and SNPs with high global missing rate were removed. This was carried out in two stages so that highly problematic SNPs, not assayed with acceptable confidence rate, and individuals do not cause systemic problems at the more stringent threshold. First, four samples and 20,307 SNPs with missing rates above 20% were removed. Second, 17 samples and 7,255 SNPs with missing rates above 5% were removed. Then, 233,882 SNPs with very low minor allele frequency (MAF), smaller than 1% in the cohort, were removed. 42,950 SNPs which grossly defy Hardy-Weinberg principle (HWE), HWE p-value smaller than 1e-10, were removed. Samples with missing rate higher than 5% on any one autosome would have been removed, but none met this criterion. Nine samples with extreme heterozygosity rates, more than 3 standard deviations from the mean rate, were removed. Six samples that were outliers on the first 10 components of multidimensional scaling, used to determine population structure, were removed. This effectively removed samples with ethnic backgrounds not captured in 1,000Genomes [58] [59] [60], or more subtle admixture. The final number of samples used from devGenes was N=239, for 295,362 SNPs.

After QC, samples were clustered based on genotype to identify population substructure of the sample. The cohort was merged with the samples from 1,000Genomes phase 3. The combined cohort was clustered into 5 groups, representing the 5 distinct super-populations found in 1,000Genomes samples. Clustering was based on the first 10 components from multi-dimensional scaling of the combined kinship matrix of the cohort and 1,000Genomes samples. The top 20 principal components of each of the 5 clusters of the cohort were calculated separately. These components were used in downstream analyses to correct for population substructure. Samples which clustered with the European 1,000Genomes samples were used for all subsequent analyses. Samples and SNPs which passed QC were imputed to the 1,000Genomes phase 3 reference using the genipe pipeline [61]. Further, imputed genotype calls were quality filtered based on the default parameters of the genipe pipeline [61]. Individual imputed genotypes with a probability less than 90% were set to missing. Imputed SNPs with more than 2% missing calls were excluded. Moreover, imputed sites with minor allele frequency in the full cohort of less 1% were excluded. LD was calculated and files were handled with PLINK [55]. Phasing of genotypes was done with SHAPEIT [62]. Imputation was performed by IMPUTE2 [63].

### 2.6 Genotyping and imputation (SPARK)

Array genotyping data was generated and processed by SPARK (Freeze three, 2019-09-12), see original publication [64] for details. SNPs were mapped to the hg19 reference of the human genome. Mapping was done with liftOver, from hg38 to hg19. All QC steps carried out with PLINK [55] and R [56], based on the QC process described in [57]. R was used for calculating standard deviations from heterozygozity and population outliers.

Samples and SNPs with missing rates above 20% were removed, then samples and SNPs with missing rates above 5% were removed. This happens in two stages so that highly problematic SNPs and individuals do not cause systemic problems at the more stringent threshold. SNPs with MAF < 1% in the cohort were removed, along with SNPs with a HWE p-value < 1e-10. Sample were removed with missing rate > 5% on any one autosome. Samples with heterozygosity rates more than 3 standard deviations from the mean rate were removed. Samples with more than 3 standard deviations from the mean on any of the first 10 MDS components were removed. Also, samples with ethnic backgrounds not captured in 1,000Genomes, or more subtle admixture were removed.

After QC, samples were clustered based on genotype to be assigned to identify population substructure of the sample. The cohort was merged with the samples from the 1,000Genomes phase 3 data. The combined cohort was clustered into 5 groups, representing the 5 distinct super-populations found in 1,000Genomes. Clustering was based on the first 10 components from multi-dimensional scaling of the combined kinship matrix of the cohort and 1,000Genomes. The top 20 principal components of each of the 5 clusters of the cohort were calculated separately. These components were used in downstream analyses to correct for population substructure. Samples which clustered with the 1,000Genomes Europeans were used for all subsequent analyses. In total, 194,410 SNPs were removed (out of 616,321 SNPs), along with 1,922 individuals removed due to QC filters.

Samples and SNPs which passed QC were imputed to the 1,000Genomes phase 3 reference using the genipe pipeline [61]. In brief, genipe performs LD calculation and file handling with PLINK [55], phasing of genotypes done with SHAPEIT [62], and imputation by IMPUTE2 [63].

### 2.7 Factor analysis

Factor analyses were performed using the factanal function in R, and fits ranging from 2 to 15 factors were attempted and characterized *post hoc* for fit and utility characteristics. For the devGenes parent report survey, a model of 11 factors was selected as a compromise between goodness-of-fit measures (which suggested a model with 10 factors was sufficient) and a jump in correlation with the masculinity measures that occurred at 11 factors (see Supplementary Figure 2). The loadings of factors 1 through 11 suggest the following interpretations: (1) academic and learning deficits, (2) sensory issues and need for order, (3) aggression, (4) feeding issues, (5) self-harm, (6) stimming, (7) social anxiety, (8) cutting, (9) low social functioning, (10) gastrointestinal issues, (11) sensitivity to sound. For the SCQ in SPARK, no fit up to 15 factors suggested a sufficient number of factors by the Chi-squared test. We selected a factor model with eight factors as a compromise between interpretability and adequate capture of variance (∼ 50%). The SCQ factors yielded from this analysis correspond approximately to joint attention (factor 1), odd behaviors (factor 2), odd speech (factor 3), nod-yes/shake-no (factor 4), appropriate facial expressions (factor 5), interest in other children (factor 6), makes conversation (factor 7), and cooperative/imaginative play (factor 8). See Figure 5B for factor loadings. We further set aside SCQ items with uniqueness > 50% (Supplementary Figure 4) for consideration separate from the factors, since they were not adequately captured by the model.

**Figure 3.**
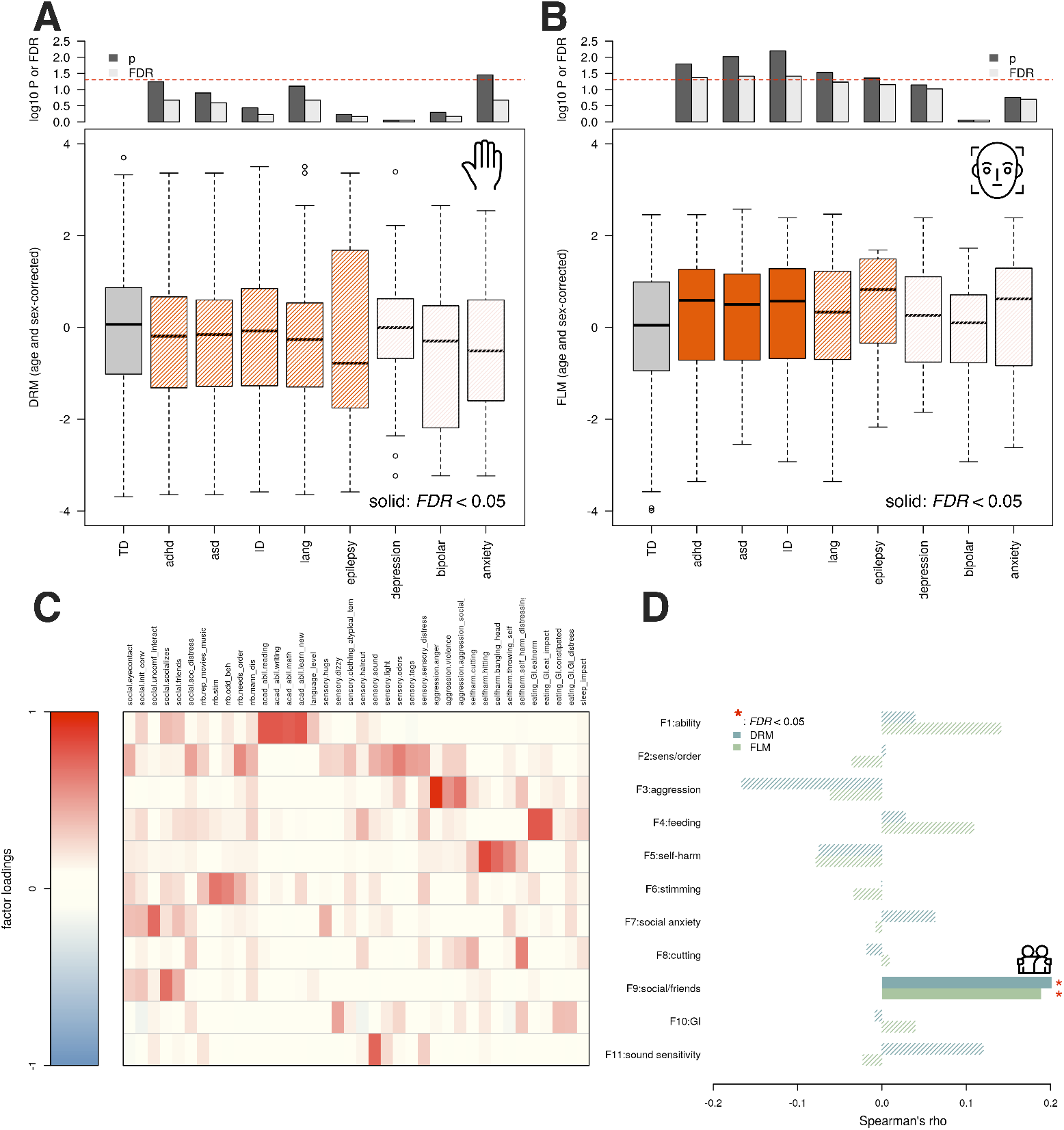
Masculinity associations with diagnoses and parent-reported problems. When comparing the digit ratio masculinity (DRM) of undiagnosed (TD) individuals and diagnostic groups, no comparison was significant after correction for multiple testing (A). In contrast, a significant positive relationship was found for facial landmark masculinity (FLM) among individuals diagnosed with ADHD, ASD, or ID (B). When examining factor scores based on parent concerns of devGenes participants across a variety of domains (C), social functioning emerged as a point of convergence, where both DRM and FLM showed positive and independent associations with a factor loading on parent-reported concerns about lack of friends and social activity (D).

**Figure 4.**
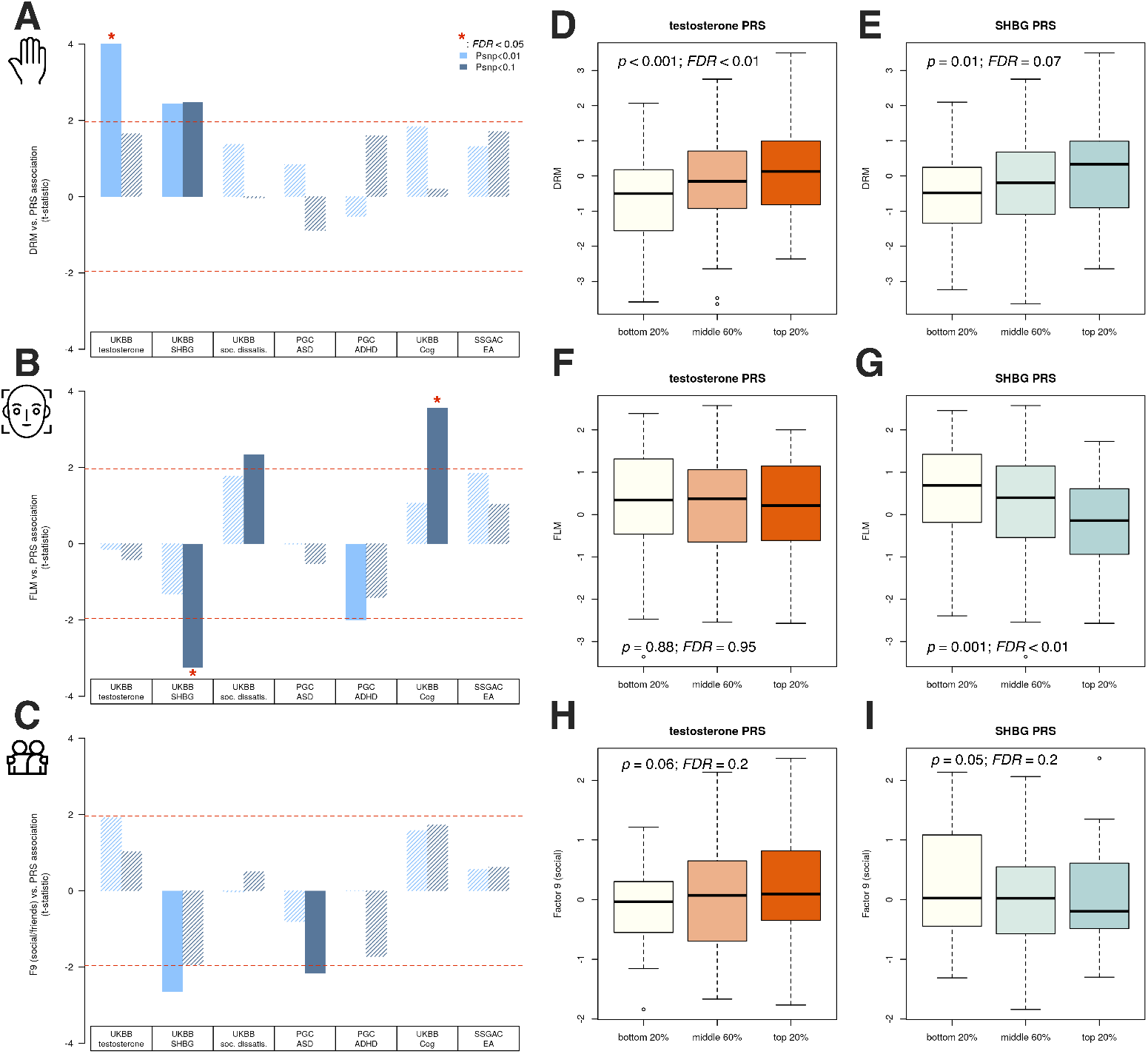
Polygenic associations with social functioning and morphological masculinity. Polygenic risk scores (PRS) for testosterone, SHBG, dissatisfaction with friendships, autism, ADHD, cognitive ability, and educational attainment were computed in the devGenes sample and used as a means to better understand potential genetic mechanisms underlying digit ratio and facial masculinity (B) (DRM and FLM, respectively), as well as the social impairment factor they predict (C) (Fig. 3D). DRM is best predicted by testosterone PRS, while FLM is best predicted by SHBG PRS (a negative relationship). The social impairment factor suggests both a positive contribution by testosterone PRS and a negative contribution by SHBG PRS (both nominally significant at *p* ∼ 0.05). The effect of testosterone and SHBG PRS (respectively) on DRM (D,E), FLM (F,G), and the social impairment factor (H,I) is shown. In (D-I), a SNP threshold of *p* < 0.01 was used for testosterone PRS and *p* < 0.1 for SHBG PRS (these are, respectively, where the associations with DRM and FLM achieved maximal significance).

**Figure 5.**
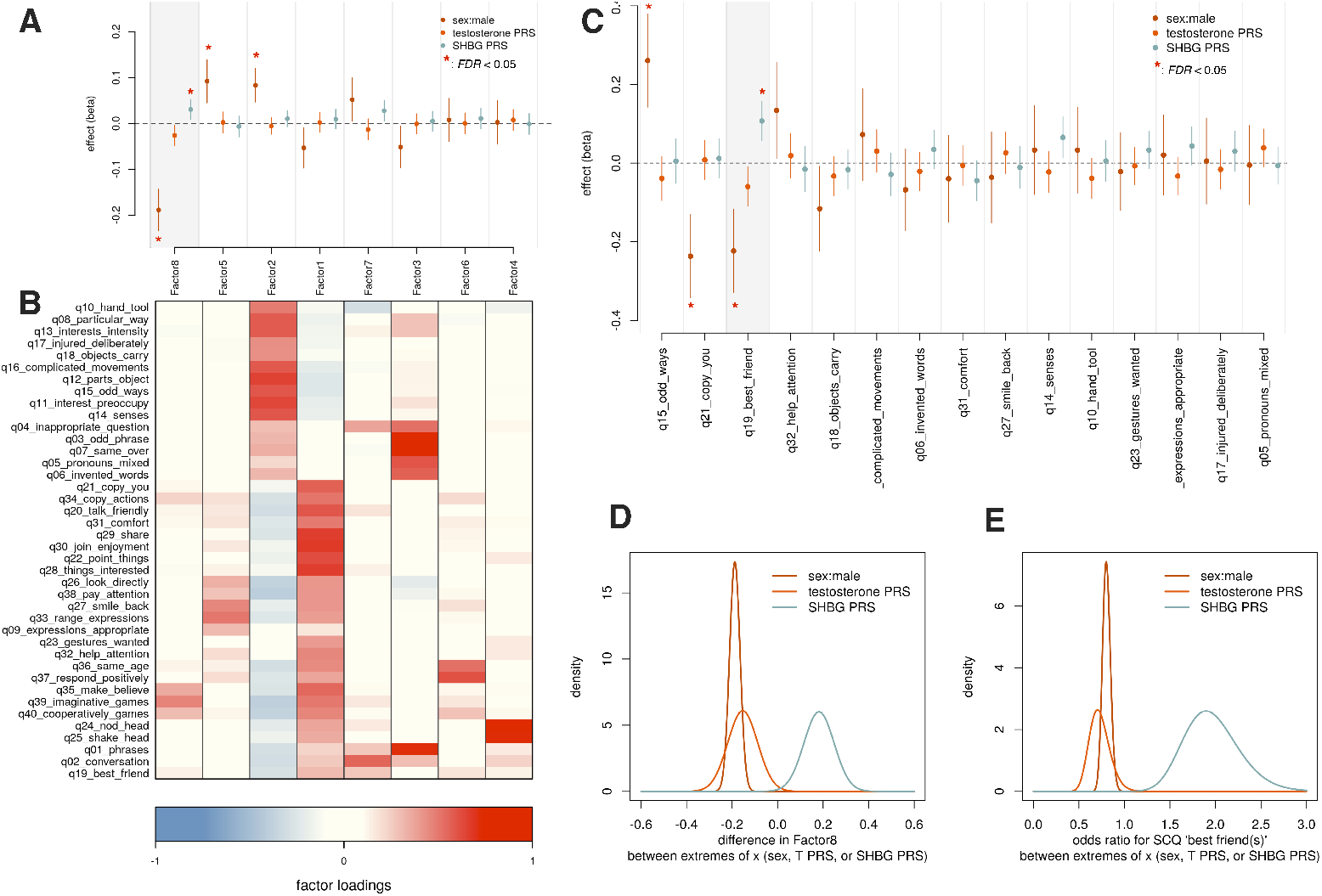
Polygenic androgen signaling associations with social functioning in SPARK. Results of the social communication questionnaire (SCQ) from N=9,419 SPARK participants were synthesized into eight latent factors, whose scores were tested for association with binary sex, testosterone PRS, and SHBG PRS (A, ordered by decreasing effect size of binary sex). Factor loadings are shown in (B). Factor 8, which loads on imaginative, creative, and cooperative play, was significantly associated (*p* < 0.05) with binary sex, testosterone PRS, and SHBG PRS, and in directions in agreement with our hypothesis. Some SCQ items were not well-captured by the factor structure, and so were considered separately (C). Of these, SCQ item 19 “has a best friend or friends” was likewise associated (*p* < 0.05) with binary sex, testosterone PRS, and SHBG PRS in the anticipated directions. When considering the extremes of each explanatory variable (sex: male, female; testosterone and SHBG PRS: 0.1 and 99.9th percentiles) and scaling the effect parameter estimates across those ranges, we observed that the binary sex contribution to Factor 8 (D) and SCQ item 19 (E) was comparable in magnitiude to the purely autosomal effects of testosterone and SHBG PRS.

### 2.8 Analysis of polygenic estimates

In order to investigate the underlying genetic factors associated with masculine features, we tested the association of masculinity (digit ratio and facial masculinity, in separate models) with polygenic risk scores relevant to autism, attention-deficit/hyperactivity disorder, educational attainment, cognitive performance, testosterone (UK Biobank field ID 30850), sex-hormone binding globulin (SHBG, UK Biobank field ID 30830), and social dissatisfaction (UK Biobank field ID 4570, here coded such that higher values correspond to increasing dissatisfaction). Briefly, a polygenic risk score (PRS) is a sum of trait-associated alleles across many genetic loci, typically weighted by effect sizes estimated from a genome-wide association study (GWAS), which can act as a biomarker for a phenotype [65]. GWAS summary statistics for the traits noted above were downloaded from the UK Biobank [66] [67] [68], Psychiatric Genomics Consortium [69] [70] [71], and Social Science Genetic Association Consortium [72]. PRS for all individuals were calculated using the PRSice tool [73]. We evaluated two SNP inclusion thresholds: *p* < 0.01 and *p* < 0.1. To test PRS-trait associations, a linear model was fit to predict the outcome with PRS as an explanatory variable (each PRS was tested independently unless otherwise stated), with sex, age, and the top five genetic principal components as covariates.

## 3 Results

### 3.1 Study 1: devGenes

#### 3.1.1 Facial landmark masculinity shows more pronounced sex and diagnostic differences than 2D:4D ratio

We examined the trends, across age, of both 2D:4D ratio and FLM within the entire (cross-sectional) devGenes cohort (Fig. 2). For 2D:4D ratio (Fig. 2A), we first examined the trends of unaffected (i.e., no reported NDD diagnosis) males and females (Fig. 2B). The 95% confidence intervals (CI) for unaffected males and females overlapped substantially until adulthood (30 years), which, although a more stringent criterion than significance testing, may suggest lower power to detect sex differences in child/adolescent samples. Comparing affected males to unaffected males (Fig. 2C), we observed that the affected trend exceeded the unaffected 95% CI for individuals in the 20s-40s age range. This suggests that affected adult males show more feminine 2D:4D ratios than their unaffected counterparts. Considering the 2D:4D ratio of affected females (Fig. 2D), we observed that the trend likewise showed a tendency toward greater femininity than unaffected females. In both cases, the null hypothesis suggested by the extreme male brain theory of *M*_*NDD*_⩽*M*_*TD*_ (where *M* is some objective measure of masculinity) could not be rejected using the digit ratio data (empirical *p* = 0.988 for males and *p* = 0.999 for females; see Supplementary Figure 4).

Turning to facial masculinity (Fig. 2E), we observed a more pronounced separation of the unaffected males and females, with complete separation of 95% CIs by age 13 years (Fig. 2F). In contrast to 2D:4D ratio, we observed that affected males appeared more masculine by facial landmarks than unaffected males, with the affected trend staying outside the 95% CI for individuals as old as 60 (Fig. 2G). Similarly, we observed that affected females displayed greater facial masculinity than their unaffected counterparts (Fig. 2H), again with the affected trend remaining outside the 95% CI even for individuals of older ages, where the sample contained fewer data points. In contrast to 2D:4D ratio, data from facial masculinity could reject the null hypothesis suggested by the extreme male brain theory of *M*_*NDD*_⩽*M*_*TD*_ (empirical *p* = 0.002 for males and *p* < 0.001 for females; see Supplementary Figure 4). Based on these observed differences across ages and sexes, all subsequent analyses used age- and sex-corrected measures of 2D:4D ratio (digit ratio masculinity or DRM) and facial landmark masculinity (FLM). We found these measures of DRM and FLM to be nominally correlated (*r* = 0.081, *p* = 0.037).

#### 3.1.2 FLM is associated with NDD diagnoses

Having become familiar with the basic patterns of 2D:4D ratio and facial masculinity in the same neurodevelopmental cohort, we next asked whether these morphological proxies for androgen exposure were significantly associated with specific diagnoses. Using t-tests, we compared the age- and sex-adjusted DRM and FLM scores in each diagnostic group (ADHD, ASD, ID, language disorder, epilepsy, depression, bipolar disorder, and anxiety disorder) with those who lacked these diagnoses (Figure 3). We found that DRM was not significantly associated with any NDD diagnoses (all *p* > 0.05) and was nominally and negatively associated with anxiety (*p* = 0.04; *t* = −2.15, *FDR* = 0.21, Fig. 3A), while FLM was significantly and positively associated with ADHD (*p* = 0.016; *t* = 2.42), ASD (*p* = 0.01, *t* = 2.6), and ID (*p* = 0.006, *t* = 2.76) even after correction for multiple testing (all *FDR* < 0.05, Fig. 3B). Language disorder and epilepsy achieved nominal significance (*p* < 0.05, *FDR* > 0.05). Taken together, these results suggest that in the devGenes cohort, increasing FLM is associated with NDDs broadly and not exclusively with any particular NDD.

#### 3.1.3 DRM and FLM are associated with parent-reported concerns about social functioning

We next turned to parent-reported concerns across a variety of domains, including social deficits, restricted and repetitive behaviors, academic performance, language, sensory issues, aggression, self-harm, eating and gastrointestinal issues, and sleep. We combined these parent reported items into latent variables using factor analysis, which yielded 11 factors (Fig. 3C, see methods). We tested the association of (age and sex-adjusted) DRM and FLM with each factor (Fig. 3D) using the Spearman correlation. The only factor for which both DRM and FLM achieved a significant association (*ρ* = 0.2, *p* = 0.004, *FDR* = 0.04 and *ρ* = 0.19, *p* = 0.004, *FDR* = 0.04, respectively) was factor 9, which loads heavily on items where parents were asked about the frequency of social interaction with individuals outside the immediate family, and a qualitative descriptor of the number of friends the child has (none, a few, many). The items related to social functioning fell into two factors, one related to social anxiety (factor 7) and the other to social functioning (factor 9). Our measures of masculinity were only significantly associated with factor 9, which may suggest some specificity of androgen-related mechanisms for social *functioning* (extent and quality of friendships and social connections) rather than social anxiety (fear of social interactions). A follow-up analysis fit a linear model predicting the factor 9 score as a function of both DRM and FLM together. Both DRM and FLM were significantly associated with factor 9 (*β* = 0.09, *SE* = 0.04, *t* = 2.15, *p* = 0.03 and *β* = 0.1, *SE* = 0.046, *t* = 2.17, *p* = 0.03), suggesting independent and additive contributions of similar magnitude.

#### 3.1.4 Polygenic association of social functioning, DRM, and FLM with testosterone and SHBG

With evidence that DRM and FLM might contribute additively to social deficits and potentially through different mechanisms, we next sought to improve our mechanistic and etiological insight into these traits by testing their association with a collection of polygenic risk scores (PRS) relevant to the topic of this investigation. We first examined parent-reported social interaction frequency and its associations with polygenic estimates of total serum testosterone (UK Biobank field ID 30850), sex hormone binding globulin (SHBG, UK Biobank field ID 30830), an item from the UK Biobank measuring satisfaction with friendships (UK Biobank field ID 4570, here coded such that higher values correspond to increasing dissatisfaction), ASD risk [71], ADHD risk [70], cognitive performance, and educational attainment [72] (Fig. 4). For DRM (Fig. 4A), we observed significant positive associations for testosterone and SHBG PRS (*p* < 0.05), though only testosterone remained significant after adjustment for multiple testing (*β* = 0.8, *SE* = 0.2, *t* = 4.0, *p* = 8.8 × 10^−5^, *FDR* = 0.001). For FLM (Fig. 4B), we observed a significant negative association for SHBG PRS and positive associations for social dissatisfaction PRS and cognitive performance PRS (all *p* < 0.05), though only SHBG (*β* = −0.58, *SE* = 0.18, *t* = −3.26, *p* = 0.001, *FDR* = 0.009) and cognitive performance (*β* = 0.69, *SE* = 0.19, *t* = 3.57, *p* = 0.0004, *FDR* = 0.006) remained significant after adjustment for multiple testing. For factor 9 (the low social functioning factor, 4C), no PRS survived multiple testing correction (*FDR* < 0.05), though importantly both testosterone and SHBG PRS met or approached nominal significance (*p* < 0.05) and in directions concordant with our working hypothesis about the net androgen exposure leading to reduced social functioning (*β* = 0.27 ± .14 and *β* = −0.41 ± 0.15 for testosterone PRS and SHBG PRS, respectively). For factor 9, there was also a nominally significant negative association with ASD PRS (*p* < 0.05), though this did not survive correction for multiple testing (*FDR* > 0.05). Using a SNP threshold of *p* < 0.01 for testosterone PRS and *p* < 0.1 for testosterone and SHBG PRS, respectively (which were the SNP thresholds where maximal association was observed in DRM and FLM), we visualized the PRS associations with each trait by binning PRS into the bottom 20%, middle 60%, and top 20% (Fig. 4D-I). This illustrates that DRM is most strongly associated with testosterone PRS, while FLM is only associated (negatively) with SHBG PRS. Social impairment (factor 9) is positively associated with testosterone PRS and negatively associated with SHBG PRS.

### 3.2 Study 2: SPARK

In study 1, results were consistent with the hypothesis that elevated testosterone and low SHBG (overall corresponding to more potent net androgen exposure) may contribute to the observed social deficits that were indexed by our morphological estimates of masculinity, DRM and FLM. In this replication study, with an order of magnitude more participants (N=9,419) than study 1, we investigated whether testosterone PRS and SHBG PRS would similarly predict indices of social functioning. We also compared the estimated effect of male sex (here a binary variable based on the presumed chromosomal sex of the participant) on these social functioning indices to the effect of testosterone PRS and SHBG PRS.

#### 3.2.1 Replication in SPARK of testosterone and SHBG association with social functioning

To test the generalization of the hypothesis that emerged from our previous analyses, that is, that higher testosterone and lower SHBG levels might lead to deficits in social behavior, we used SPARK, a large, US-based nationwide genetic study of autism spectrum disorders. In addition to genetic data, SPARK has a host of phenotypic data relevant to ASD. Given our working hypothesis relating to social deficits, we focused on the results of the Social Communication Questionnaire (SCQ) [46], which has 40 items designed to ascertain social communication issues typically seen in ASD. The SCQ was originally devised as a screen to use in binary clinical decision-making, though it has been used extensively in research. We again used factor analysis (see methods) to decompose the SCQ items into latent variables (eight factors, Fig. 5A-B). In addition, we performed item-level tests of association for SCQ items that were poorly captured in the factor model (see methods, Fig. 5C). Using bivariate linear or logistic regression models (polygenic estimates of testosterone and SHBG as explanatory variables), we tested association of PRS with indices of social function. Among the factors, only factor 8, which loads on items relating to cooperative and imaginative play, showed significant associations with male sex(*β* = −0.19, *SE* = 0.02, *t* = −8.15, *p* = 4.18 × 10^−16^, *FDR* < 0.01), testosterone PRS(*β* = −0.03, *SE* = 0.01, *t* = −2.34, *p* = 0.02, *FDR* = 0.16), and SHBG PRS(*β* = 0.03, *SE* = 0.01, *t* = 2.74, *p* = 0.006, *FDR* = 0.049). Notably, all associations were in the direction predicted by our working hypothesis (note that in this case, a higher score for factor8 indicates more prosocial behavior). Among the individual items not well-captured by the factor model (Fig. 5C), only SCQ item 19 (“has a best friend or friends”) was significantly associated with male sex (*β* = −0.22, *SE* = 0.05, *Z* = −4.15, *p* = 3.27 × 10^−5^, *FDR* < 0.01),testosterone PRS (*β* = −0.06, *SE* = 0.03, *Z* = −2.31, *p* = 0.02, *FDR* = 0.31), and SHBG PRS(*β* = 0.11, *SE* = 0.03, *Z* = 4.16, *p* = 3.15 × 10^−5^, *FDR* < 0.01). Again, the directions of these associations were in agreement with our working hypothesis (note that again, the direction is such that a higher value indicates prosocial behavior).

#### 3.2.2 Comparing effect of binary sex to autosomal polygenic sex hormone effects

The effects noted for testosterone and SHBG PRS in the above linear and logistic regression models are on a “per-SD of PRS scale”. That is, for each increase of one standard deviation of PRS, there will be an expected increase (or decrease) in the trait of *β* ± *SE*. To meaningfully compare these PRS estimates to that of binary sex, which is an effect from one extreme of the variable (female or 0) to the other (male or 1), we must scale the PRS effects to likewise span their extremes. In this case, we choose the PRS values at the 0.1%-ile and the 99.9%-ile. The difference between these PRS values (which have already been scaled to mean 0 and SD 1) acts as a scaling factor for both the PRS *β* and its standard error. Using this approach, we find that the expected effect of binary male sex is comparable or even perhaps less extreme than the apparent effects of testosterone and SHBG PRS (factor 8: male sex: −0.19 ± 0.03, testosterone PRS: −0.15 ± 0.066, SHBG PRS: 0.18 ± 0.066, see Fig. 5D; SCQ item 19: male sex:−0.22 ± 0.05 testosterone PRS: −0.35 ± 0.15, SHBG PRS: 0.64 ± 0.15, see Fig. 5E).

## 4 Discussion

The current study employed a two-step approach to investigate genetic factors associated with both digital and facial masculinity and their connection to neurodevelopmental outcomes. In the first study, which was the first of its kind to examine both digit ratio and facial masculinity in the same genetically characterized neurodevelopmental cohort, we found convergent evidence that androgen exposure is associated with deficits in social functioning, even while correcting for binary sex. We found that facial landmark masculinity (FLM) was predictive of NDD diagnoses (ADHD, ASD, ID), in the direction consistent with the extreme male brain theory, while digit ratio masculinity (DRM) was not. The predictive power of FLM relative to DRM suggests that further pursuit of biomarkers and endophenotypes based on facial morphology may prove more fruitful than digit ratio. Particularly noteworthy is that social functioning showed convergent association with both FLM and DRM. Through analysis in connection with polygenic risk scores (PRS), our findings implicate both testosterone and sex hormone binding globulin (SHBG) levels as complementary and potentially additive factors that may impact social functioning. In the second study, our hypothesis found additional statistical support in the large SPARK cohort, and we found evidence suggesting that the autosomal genetic factors (i.e., the PRS) that predict testosterone and SHBG levels may exert effects on social functioning that are comparable in magnitude to the effect of binary (i.e., chromosomal) sex itself. This provides evidence that, at least in the context of its effect on social functioning, sex may be more accurately described as bimodal and continuous, rather than binary and discrete.

Notably, the direction of associations between FLM and NDD diagnosis was consistent with the extreme male brain theory (i.e., greater masculinity conferred greater risk for NDD traits), which was not the case for DRM. Although DRM was associated with social deficits in the expected direction, when considering clinical diagnoses, young-to middle-adulthood males with NDDs demonstrated more *feminine* digit ratios than their typically-developing peers. This is not the first study to find that androgynous features are associated with NDDs [33] and that males with ASD demonstrate more feminine digit ratios [36]. However, quantitative reviews have supported the association between masculine digit ratio and ASD [74] [75], so the literature on DRM remains mixed. It is also unclear why the association between DRM and affected status did not emerge until young-adulthood, given that NDD-related outcomes emerge in early childhood. Longitudinal studies of typically developing children show that digit ratio tends to increase (i.e., becomes more feminine) across puberty [76] [77], however the rank order of inter-individual differences remain stable. No study to date has examined the developmental trajectory of DRM in individuals with NDDs, or longitudinally compared groups of individuals with and without NDDs, so further research in this area may be useful. Nevertheless, our results suggest that FLM may provide greater utility than DRM as a biomarker with clinical relevance.

The differential performance between FLM and DRM in predicting NDD traits may reflect a number of physiological factors that differentiate the two measures of masculinity. First, combining distances between 12 facial landmarks naturally provides a richer basis for an estimator of masculinity than the average length of two digits. As such, the face inherently provides a more sensitive indicator for subtle differences in biological risk. In contrast to digits, faces are more proximal to the brain not just physically, but in development and biology. Prenatal brain development is intimately tied to craniofacial development through a combination of biochemical mechanisms and genetic signaling [29]. Brain and craniofacial tissues originate from the same population of cells, such as neural crest cells, which differentiate early in fetal development to eventually form the brain and face [78]. As such, developmental perturbations in the brain are more likely to manifest in craniofacial features than digits [79] [80] [81]. These developmental mechanisms support our findings that FLM is associated with NDDs broadly, and not only to ASD. Given the ties between neurodevelopment and craniofacial development, it follows that facial morphology is a useful basis for predicting neurodevelopmental risk generally, rather than ASD in particular. This was underscored by the convergence of DRM and FLM on a factor that loads on social functioning. That this convergence on social impairment was found in a mixed NDD cohort is noteworthy: although ASD is most prominently characterized by deficits in social functioning, other NDDs are also marked by elevated rates of social dysfunction [82] [83], underscoring the fact that key features of NDDs, and psychopathology more broadly, do not adhere to diagnostic boundaries [43] [44].

To provide potential mechanistic insights, we investigated the association of specific polygenic risk scores to DRM, FLM, and factor 9, a latent variable capturing social impairment which was found to be a point of convergence between FLM and DRM. Both DRM and FLM were significantly predicted by androgen-related PRS: DRM by testosterone and FLM by SHBG, an attenuator of free testosterone. These and other results of our investigations suggest that DRM and FLM may access a latent masculinity through complementary androgen-related mechanisms. Social impairment (factor 9) showed a combined nominal association where increasing testosterone PRS and decreasing SHBG PRS were independently and additively associated with increased social deficits. Taken together, these results pointed toward a hypothesis of increased net androgen exposure resulting in reduced social functioning.

Our investigation in the much larger SPARK cohort (N=9,419) added further evidence in favor of this hypothesis, as male sex, testosterone PRS, and SHBG PRS were all significantly associated, in the anticipated direction, with cooperative/imaginative play, and having a best friend or friends. It is noteworthy that these indicators emerged from a background of other possible social phenotypes, including joint attention, facial expressions, and binary communication (yes/no by head nodding or shaking). Our working hypothesis was based on the initial observation of a testosterone/SHBG association with a factor that, as in the SPARK study, loaded on friendships and social interactions (and not on other social phenotypes such as social anxiety). Further investigation is needed, but the fact that both the discovery sample and the replication sample converged on a friendship-oriented social phenotype is striking.

When considering the genomic reservoir of androgen-related risk, it seems intuitive that the sex chromosomes (and therefore our binary concept of male and female sex) would play a dominant role. However, our comparison of sex chromosome and autosomal effects (binary sex and polygenic estimates of testosterone and SHBG, respectively, see Fig. 5D,E) led to the unexpected finding that sex chromosome and autosomal contributions are comparable to each other in magnitude, with respect to risk of social deficits. Although perhaps not intuitive, this should not be surprising, given that a sizable majority of genes involved in androgen metabolism and sex differentiation pathways are encoded on autosomes (see Supplementary Table 5), and that genetic variation in these genes will shape and tune an individual’s response to the presence of sex hormones throughout their development. While further research is needed, these findings suggest the possibility of a latent and continuous “autosomal sex” that, like binary sex, plays a major role in neurodevelopmental outcomes, especially those related to social functioning.

How might the proposed genetic risk mechanisms impact social functioning? Sex hormones affect brain structure and function through multiple mechanisms and on different time scales. Human and animal research suggests that both short-term (e.g., neurotransmitter and neuropeptide) and long-term (i.e., developmental) mechanisms downstream of androgen exposure can lead to reduced capacity for social functioning [84] [85] [86] [87]. Studies in humans that have linked fetal testosterone exposure (FTE) to later reduced quality of social relationships [88] and reduced empathic descriptions of social interactions [89] underscore the long-term processes, potentially related to neural circuit formation, that may explain androgen-related social deficits. Taken together, the early predictive value of FTE and our observation that affected individuals show elevated masculinity relative to typically developing peers, even at an early age (Fig 2. panels G,H), suggest that indices of sex hormone exposure may hold promise as early markers of developmental risk. The more immediate behavioral effects of androgens might be explained by their interactions with signaling pathways involving the neuropeptides oxytocin and vasopressin, which have been well-established as key mechanisms in social behavior [90] [91], and have recently shown promise in clinical trials for their influence on social functioning [92]. Our results build on this previous work by providing genetic evidence that increasing testosterone and decreasing SHBG (overall corresponding to more potent net androgen exposure) may underlie the observed social deficits that were indexed by our morphological estimates of masculinity, DRM and FLM. Furthermore, our results may offer a partial explanation for why males are diagnosed with NDDs at high rates than females [1]: males, on average, will have higher androgen exposure and production throughout their lives, and are consequently at greater risk for disorders where social impairment is a factor in diagnosis, such as ASD and other NDDs. Conversely, females have lower androgen production and exposure throughout their lives, making this risk mechanism generally less applicable and resulting in a lower rate of diagnosis with disorders where social impairment is a decisive factor. A key contribution of this work is that we show that *autosomal* androgen-related risk mechanisms (shown here as polygenic risk scores) can and do operate in males and females alike, contributing risk above and beyond that linked to sex chromosomes.

## 5 Limitations

Despite the replication of our findings, our study has several limitations that are important to acknowledge when considering the trajectory of future work. First, we did not directly measure sex hormones, but instead used well-supported upstream (polygenic estimators) and downstream (morphological features) correlates as proxies for androgen exposure. While these are convenient to measure in large samples, further testing and refinement of hypotheses connecting genetics, sex hormones, and social functioning will require direct measurement of the relevant factors. Next, because our study was not interventional, the precise structure of causal relationships between sex hormones and social functioning remains unclear. Studies of short-term testosterone exposure have demonstrated significant effects on some behaviors such as risk taking [93] and aggression [94] but not others, such as empathizing [95]. Further interventional research across a range of exposure durations, for example in samples of patients undergoing hormone therapy [96], will be needed to further elucidate the mechanisms underlying the link between androgens and social functioning.

## Data Availability

Basic individual-level data for devGenes participants is included in Supplementary Table 3, and we have made code available at https://research-git.uiowa.edu/michaelson-lab-public/facial-masculinity-2021 to reproduce the analyses presented in figures. Data from the SPARK cohort are made available to researchers through https://base.sfari.org.

## 6 Declarations

### 6.1 Ethics approval and consent to participate

Study procedures were approved by the University of Iowa’s Institutional Review Board, and informed consent was obtained for each participant (IRB 201505743).

### 6.2 Consent for publication

Assent was obtained from the subject and consent from the legal guardian for the likeness appearing in Figure 2.

### 6.3 Availability of data and materials

Basic de-identified individual-level data pertaining to the manuscript findings are available in Supplementary Table 3. Code to reproduce our figures and analyses are available at https://research-git.uiowa.edu/michaelson-lab-public/facial-masculinity-2021. Please contact the authors for all other data requests.

### 6.6 Authors’ contributions

BGM coordinated the recruitment of participants, organized data, and wrote the manuscript. YH led the analyses and wrote the manuscript. KV contributed to analyses and wrote the manuscript. DH, MC, SAM, FL, AZ, JYK, ST, LB, EB, TK, NP, TK, LC, and TRT contributed to recruitment activities, data organization, and analyses. JYK and ST managed participant biospecimens. JJM and TK prepared code for public release. JJM led the study, performed analyses, and wrote the manuscript. All authors read and approved the final manuscript.

## 6.7 Acknowledgements

We would like to thank all devGenes participants for their important contributions to this research. We would also like to thank Atticus Michaelson for his permission to use his image in Figure 2. We are grateful to all of the families in SPARK, the SPARK clinical sites and SPARK staff. We appreciate obtaining access to genetic and phenotypic data for SPARK data on SFARI Base. Approved researchers can obtain the SPARK population dataset described in this study by applying at https://base.sfari.org.

